# Mapping the distribution of zero-dose children to assess the performance of vaccine delivery strategies and their relationships with measles incidence in Nigeria

**DOI:** 10.1101/2022.10.10.22280894

**Authors:** C. Edson Utazi, Justice M. K. Aheto, Adelle Wigley, Natalia Tejedor-Garavito, Amy Bonnie, Christopher C. Nnanatu, John Wagai, Cheryl Williams, Hamidreza Setayesh, Andrew J. Tatem, Felicity T. Cutts

## Abstract

Geographically precise identification and targeting of populations at risk of vaccine-preventable diseases has gained renewed attention within the global health community over the last few years. District level estimates of vaccination coverage and corresponding zero-dose prevalence constitute a potentially useful evidence base to evaluate the performance of vaccination strategies. These estimates are also valuable for identifying missed communities, hence enabling targeted interventions and better resource allocation. Here, we fit Bayesian geostatistical models to map the routine coverage of the first doses of diphtheria-tetanus-pertussis vaccine (DTP1) and measles-containing vaccine (MCV1) and corresponding zero-dose estimates at 1×1 km resolution and the district level using geospatial data sets. We also map MCV1 coverage before and after the 2019 measles vaccination campaign in the northern states to further explore variations in routine vaccine coverage and to evaluate the effectiveness of both routine immunization (RI) and campaigns in reaching zero-dose children. Additionally, we map the spatial distributions of reported measles cases during 2018 to 2020 and explore their relationships with MCV zero-dose prevalence to highlight the public health implications of varying performance of vaccination strategies across the country. Our analysis revealed strong similarities between the spatial distributions of DTP and MCV zero dose prevalence, with districts with the highest prevalence concentrated mostly in the northwest and the northeast, but also in other areas such as Lagos state and the Federal Capital Territory. Although the 2019 campaign reduced MCV zero-dose prevalence substantially in the north, pockets of vulnerabilities remained in areas that had among the highest prevalence prior to the campaign. Importantly, we found strong correlations between measles case counts and MCV RI zero-dose estimates, which provides a strong indication that measles incidence in the country is mostly affected by RI coverage. Our analyses reveal an urgent and highly significant need to strengthen the country’s RI program as a longer-term measure for disease control, whilst ensuring effective campaigns in the short term.

**Highlights:**

- In 2018, about 8,883,931 and 12,566,478 children aged under 5 years old in Nigeria had not received routine DTP1 and MCV1, respectively.
- MCV and DTP zero-dose prevalence shared similar patterns, with highest prevalence districts concentrated in the northeast and northwest but also found elsewhere
- Measles incidence, though mitigated by campaigns, is related to subnational routine MCV1 coverage
- Residual zero-dose prevalence following vaccination campaigns should be analysed and reported
- Targeted campaigns or routine immunization interventions in higher zero-dose prevalence districts are needed to reduce disease risk

## Introduction

Achieving high rates of vaccination coverage is vital for disease control, elimination and eradication. Since the introduction of the WHO Expanded Programme on Immunization (EPI) in 1974, substantial gains in vaccination coverage have been made globally through a combination of routine immunization (RI) and campaign strategies. However, within the last decade, coverage levels have stalled or regressed in many countries [1–3]. This situation is further exacerbated by the ongoing COVID-19 pandemic, which has caused severe disruptions to vaccination services globally, leading to postponement of campaigns and inadequate routine immunization service delivery [4–6]. In 2020, it was estimated that 22.7 million children missed out on routine immunization – a 19.5% increase from 2019, with the number of children who had received no vaccines increasing from 13.6 million in 2019 to 17.1 million in 2020 [5]. This implies that countries, districts and communities where these un- and under-immunized populations reside continue to be at risk of vaccine-preventable diseases (VPDs).

Inequities in vaccination coverage and vulnerabilities to VPDs most often occur because of suboptimal RI performance and/or ineffective vaccination campaigns. To identify these vulnerable populations, spatially detailed data are required, beyond the large-area summaries reported from household surveys. Such data are crucial for mapping areas that are un- or under-vaccinated via each delivery method [7, 8] and understanding which strategies to adopt to fill gaps and boost coverage. Geospatial analyses have now gained traction as a vital tool for creating high-resolution and district level maps of health and demographic indicators [1, 7–11]. In the case of vaccination coverage, these maps are integrated with relevant gridded population data to produce estimates of numbers of un-vaccinated (i.e., zero-dose prevalence) and under-vaccinated populations at various spatial scales, thus helping with identifying and delineating clusters of vulnerabilities within countries and better allocation of resources. Such spatially detailed data also enable integration with other data sources such as health facility catchment maps and locations of vaccination posts, to give more complete health metrics or decision-making information. The programmatic relevance of spatially detailed data for immunization is well recognized by global health policy frameworks such as the WHO Immunization Agenda 2030 [12] which has a target of achieving a 50% reduction in numbers of “zero-dose” children by 2030, and Gavi Strategy 5.0 [13] which aims to achieve equity in vaccination coverage and a 25% reduction in the number of zero-dose children by 2025 through reaching missed communities. Furthermore, the distribution of zero-dose populations, when combined with disease incidence, could present a fuller picture to evaluate the relative performance of RI and campaigns. Where disease surveillance systems have consistent reporting rates over time [14], decreases in both reported incidence and zero-dose prevalence following a campaign are a more compelling demonstration of the impact of the campaign. Also, hotspots of susceptibility as evidenced by high zero-dose prevalence and high incidence, are most likely indicative of poor RI performance, suboptimal campaigns or the failure of RI to sustain and improve upon gains made through campaigns, particularly in high birth rate settings.

In 2019 and 2020, Nigeria was identified as being among the top 3 countries with the most un- or under-vaccinated children globally [2]. WHO and UNICEF estimates of national immunization coverage (WUENIC) show that the coverage of basic vaccines such as DTP3 and MCV1 has not improved much in recent years, standing at 57% and 54% respectively in 2020 [2]. Geospatial analyses of data from various surveys conducted in the country since 2013 [1, 8, 10, 15] have shown a persistent north-south divide in coverage, with the northern states having relatively low coverage levels despite concerted efforts to improve coverage levels across the country. A recent analysis of measles case-based surveillance data also found higher incidence rates in the north, in addition to a high proportion of MCV zero-dose individuals (70.8%) among confirmed cases during 2008-2018 [16]. Several studies have identified different demand- and supply-side factors, such as maternal access to and utilization of health services, maternal education, religion, ethnicity, wealth, maternal age, mobile phone usage, poor attitude of health workers and vaccine stockouts as being responsible for the slow rate of progress within the country [17–19]. All of this points to an urgent need to identify and prioritize high-risk areas for effective follow up through appropriate routine and campaign strategies and robust disease surveillance[14], to put the country on a path to achieving its disease control and elimination targets.

Our subnational assessments of the relative effectiveness of RI and campaigns have typically focused on comparing maps of DTP3 and MCV1 coverage [7] or analyzing coverage maps of post-campaign coverage survey (PCCS) indicators [8]. Here, we focus on the subnational distributions of DTP and MCV zero-dose children. We use the term ‘zero-dose’ to refer independently to non-receipt of DTP (i.e., DTP zero-dose) and non-receipt of MCV (i.e., MCV zero-dose) vaccines. Specifically, we examine the performance of RI in 2017-2018 using the spatial distributions of DTP and MCV zero-dose estimates produced using the 2018 Nigeria Demographic and Health Survey (NDHS). We also assess the specific and combined performance of RI and the 2019 measles campaign (northern states only) using MCV zero-dose estimates produced through mapping MCV1 coverage before and after the campaign using the 2019 PCCS. Finally, we triangulate the zero-dose estimates with aggregate measles case-based surveillance data during 2018 – 2020 to explore the spatial relationships between RI and post-campaign MCV zero-dose prevalence and measles incidence.

## Methods

### Vaccination coverage data from the 2018 NDHS and 2019 PCCS

Routine immunization coverage data for DTP1 and MCV1 were obtained from the 2018 NDHS for children aged 12-23 months [20]. The 2018 NDHS used a stratified, two-stage sampling design to produce estimates of health and demographic indicators, including vaccination coverage, at the national, regional and state levels and for urban and rural areas. Stratification was achieved by separating each of the 36 states and the Federal Capital Territory (FCT) into urban and rural areas. Samples were drawn from within each stratum in two stages: the first stage involved the selection of survey clusters (enumeration areas) from a national sampling frame using a probability proportional to size sampling scheme, while the second stage involved selecting households randomly from household lists within the selected clusters. In all, the survey was implemented in a total of 1389 clusters, with 11 of the 1400 clusters selected initially dropped due to security reasons. Fieldwork took place between August and December 2018. This was within one year of completion of the 2017-18 national follow-up measles vaccination campaign targeting children aged 9 to 59 months [20], which was conducted during October - December 2017 in the northern states and February - March 2018 in the southern states [8]. As discussed later, there could be some misclassification of campaign doses as RI for children without documentation of RI vaccination hence we consider our estimate of routine MCV1 coverage for children aged 12-23 months “an upper bound estimate”.

For each vaccine, we used information obtained from both home-based records and maternal/caregiver recall. Hence, our analysis captures crude estimates of coverage [21]. At the cluster level, we aggregated the individual level data to produce numbers of children surveyed, numbers vaccinated and empirical proportions of children vaccinated.

The 2019 measles campaign in Nigeria was conducted between September and December 2019 in the 20 northern states only. Post-campaign coverage surveys (PCCS) were implemented in each state within two weeks of conclusion of the campaigns. However, at the time of analysis, data were not available for Niger and Kogi states, leaving only 18 states for the analysis. Information on receipt of MCV1 was based on campaign cards or maternal/caregiver recall, and data were collected for all eligible children aged 9-59 months. However, our analysis is restricted to children aged 9-35 months to exclude those that may have participated in the previous campaign. We followed the methodology implemented in a previous analysis and extracted data for six PCCS indicators [8], although we report here only *MCV coverage before the campaign* and *coverage with at least one dose of MCV* by the end of the campaign.

All the extracted cluster-level vaccination coverage data are displayed in Figure 1, showing where lower and higher coverage levels were observed at the cluster level for each indicator.

**Figure 1:**
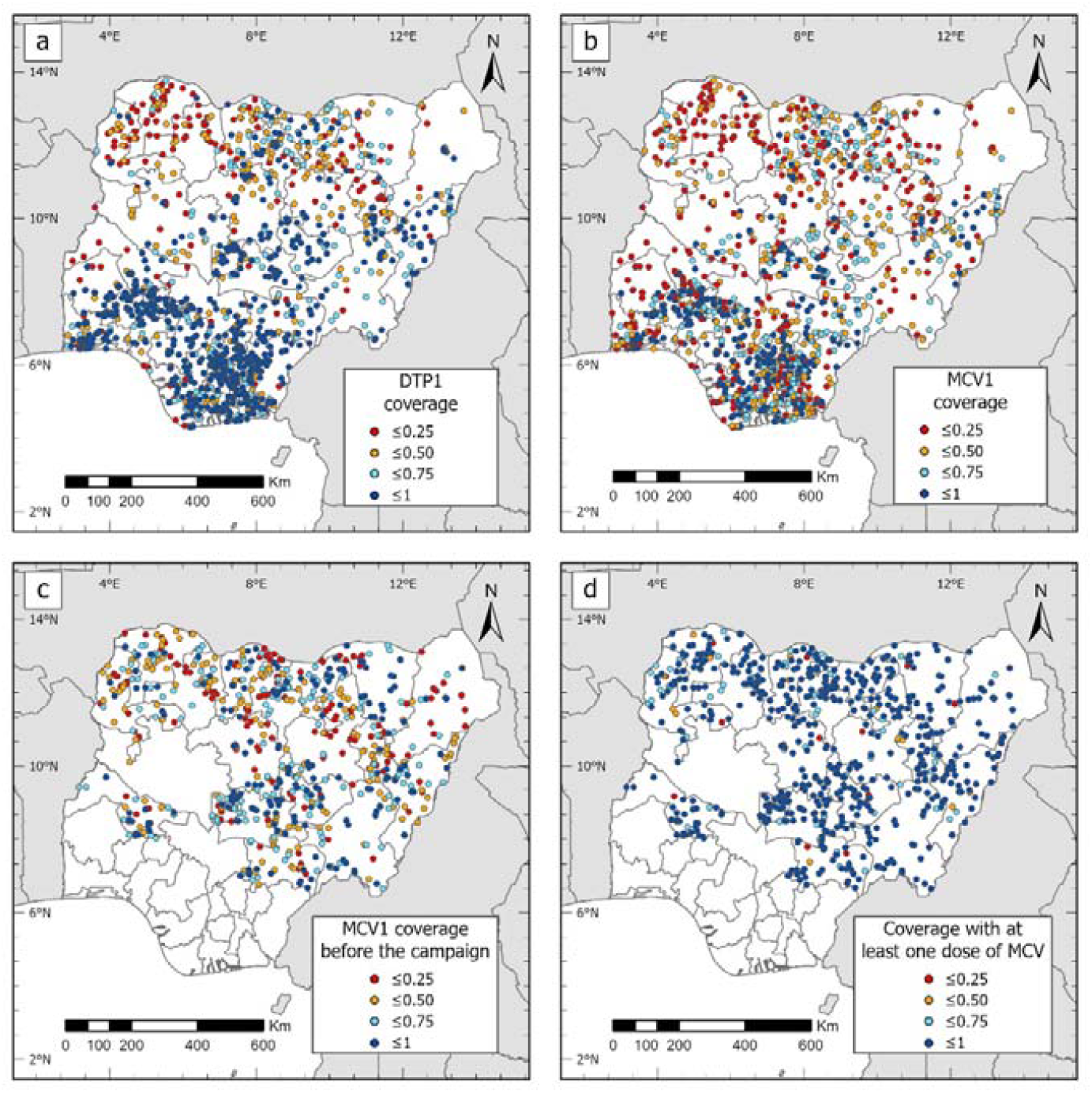
Cluster-level maps of MCV1 and DTP1 coverage (a and b, from DHS), and MCV1 coverage before the campaign and coverage with at least one dose of MCV by the end of the campaign (c and d, from 2019 PCCS)

### Geospatial covariate data, covariate selection and population data

Geospatial covariates are essential in geostatistical modelling to explain and predict the outcome variable, although the latter rationale is paramount. As in previous work [7, 8, 10], we assembled a suite of geospatial socio-economic, environmental, and physical covariates for the analyses. These included travel time to the nearest health facility (providing routine immunization services), poverty, economic index, nightlight intensity, livestock density, distance to conflicts and land surface temperature (see [7, 8, 10]). These covariates were processed as detailed in previous work to produce 1×1 km raster data sets and cluster level data using the geographical coordinates from each of the surveys.

Following previous work [7, 8, 10], covariate selection was carried out to determine the best subset of covariates for modelling each indicator. The covariate selection process involved checking the relationships between the covariates and (empirical logit transform of) vaccination coverage and applying the log transformation where necessary to improve linearity; fitting of single covariate models and ranking the covariates based on their predictive ability (i.e. using predictive R^2^ values); checking for multicollinearity and selecting between highly correlated covariates (correlation > 0.8 or variance inflation factor > 4.0) using their ranks; and using stepwise regression (backward elimination based on Akaike Information Criterion (AIC)) to choose the best model/combination of covariates for modelling the indicator in a non-spatial framework using binomial regression models. For the analysis using PCCS data, we additionally created a uniform set of covariates for the modelled outcome indicators – see Utazi et al [8].

To enable the production of coverage estimates for different administrative areas, as is required in geospatial estimation of health and demographic indicators, we obtained population estimates for children aged under 5 years from WorldPop (www.worldpop.org) [22]. The data were also used to produce estimates of numbers of children under 5 years who had not received DTP1 or MCV1, otherwise known as DTP and MCV zero-dose children.

### Measles case-based surveillance data

Laboratory-supported measles surveillance has been in place in Nigeria since 2006 [16]. Routine measles case-based surveillance data are collected at the health facility level and then transmitted to the district (or local government area (LGA)), state and national levels. As recommended by WHO, suspected cases of measles are classified as confirmed by one of: a laboratory assay (Immunoglobulin M (IgM)) positive for measles, an epidemiologic linkage to a laboratory-confirmed case, or clinical signs and symptoms meeting the measles clinical case definition [23]. The data were obtained for the years 2018 to 2020 and then summarized at both the district and state levels. Relevant variables included in the data for each suspected case were date of birth/age, sex, vaccination status/number of doses of MCV received, case address (ward, district and state), date of onset of illness, urban/rural, final classification (laboratory confirmed or laboratory-discarded, epidemiological linkage, or clinically compatible) and outcome (survived or died). The confirmed measles case counts (i.e., excluding discarded cases) are displayed in Figure 4. Other summaries of the data are included in supplementary materials.

### Geospatial model fitting, validation and prediction

To model and predict vaccination coverage at 1×1 km resolution, we fitted geostatistical models with binomial likelihoods. For *i=* 1, *…, n* and a given indicator, where *n* is the number of survey locations, let *Y*(***s***_*i*_) denote the number of children vaccinated at survey location ***s***_*i*_ and *m*(***s***_*i*_) the number of children sampled at the location. The first level of the model assumes that

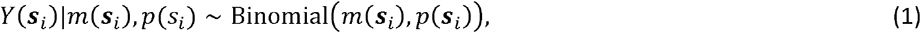

where *p*(***s***_*i*_) (*0* ≤ *p*(*s*_*i*_) ≤ **1**) is the true vaccination coverage at location ***s***_*i*_. We model *p*(*s*_*i*_) using the logistic regression model as

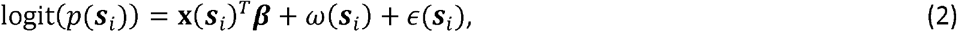

where **x**(***s***_*i*_) is the vector of covariate data associated with ***s***_*i*_, ***β*** is a vector of the corresponding regression coefficients, *E*(***s***_*i*_) is an independent and identically distributed (iid) Gaussian random effect with variance, 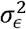, used to model non-spatial residual variation, and ω (***s***_*i*_) is a Gaussian spatial random effect used to capture residual spatial correlation in the model. That is, **ω***=* (ω (***s***_*1*_),*…, ω* (***s***_*n*_))^*T*^ *~ N*(*0*, Σ _*ω*_). Σ *ω* is assumed to follow the Matérn covariance function [24] given by 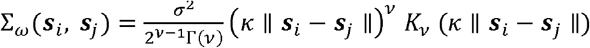, where II. II denotes the Euclidean distance between cluster locations ***s***_*i*_ and ***s***_*j*_, *σ*^2^ *> 0* is the marginal variance of the spatial process, κ is a scaling parameter related to the range 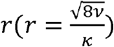 – the distance at which spatial correlation is close to 0.1, and *K ν* is the modified Bessel function of the second kind and order *ν>* 0. Further, for identifiability reasons, we set the smoothing parameter, *v= 1*, see [25].

For the PCCS indicators – *MCV1 coverage before the campaign* and *coverage with at least one dose of MCV by the end of the campaign* - we adopted the conditional probability modelling framework described in [8] to ensure that the modelled estimates were consistent with those of other indicators. We also note that a similar approach proposed in [26] could be used for estimating the coverage of the DTP dose series, but we did not consider this necessary here as we were only interested in the coverage of the first dose, DTP1.

In each case, the model described in equations (1) and (2) was fitted in a Bayesian framework using the integrated nested Laplace approximation – stochastic partial differential equation (INLA-SPDE) approach (INLA-SPDE) approach [25, 27]. Using the fitted models, we obtained predictions at 1×1 km resolution. Further, using the posterior samples of the 1×1 km predictions, we obtained the district and state level predictions as population-weighted averages taken over the 1×1 km grid cells falling withing each district or state.

Methods for assessing the out-of-sample predictive performance of the fitted model are detailed elsewhere [8–10]. Here, we rather focus on describing the patterns in the modelled estimates. All analyses were carried out in R [28] and through using the R-INLA package [29].

## Results

### DTP1 and MCV1 Coverage maps and zero-dose estimates

In Figure 2(a), we present the DTP1 and MCV1 coverage estimates (including documented and verbal recall evidence of vaccination) at 1×1 km and the district and state levels to examine routine vaccination coverage in children aged 12-23 months in 2018. The corresponding uncertainty estimates are presented in the supplementary Figure 1. The patterns in the routine coverage of both vaccines are very similar, although MCV1 coverage estimates are generally lower than DTP1 coverage estimates as expected, due to the dropouts that often occur between the two vaccines. There are visible spots of higher coverage in more urban areas, especially in the southern regions (see Aheto et al [18]), but MCV1 coverage appears more heterogenous in the south compared to DTP1 coverage. Supplementary Figure 2 shows that the dropout rates between both vaccines (relative to DTP1 coverage) varied substantially across the country. Areas with the highest positive dropout rates are spread across the six regions (e.g., Taraba, Plateau, Cross River, Akwa Ibom, Kogi and Oyo states); whereas areas with the most negative dropout rates (i.e., MCV1 coverage was higher than DTP1) are located mostly in the northwest, the northeast and some coastal areas of the south-south region (Bayelsa and Delta states). Interestingly, some of the areas with the highest positive dropout rates were also areas where higher DTP1 coverage was estimated (e.g., Akwa Ibom, Cross-River and Plateau states). Also, as was shown in previous studies [7, 8, 10], there is an apparent north-south divide in the routine coverage of both vaccines, with poorer coverage levels more pronounced in the northwest and northeast. At the district level, the lowest coverage areas (≤20%) are concentrated in Sokoto and Zamfara states for both vaccines. Both states also have the lowest coverage rates for both vaccines.

**Figure 2:**
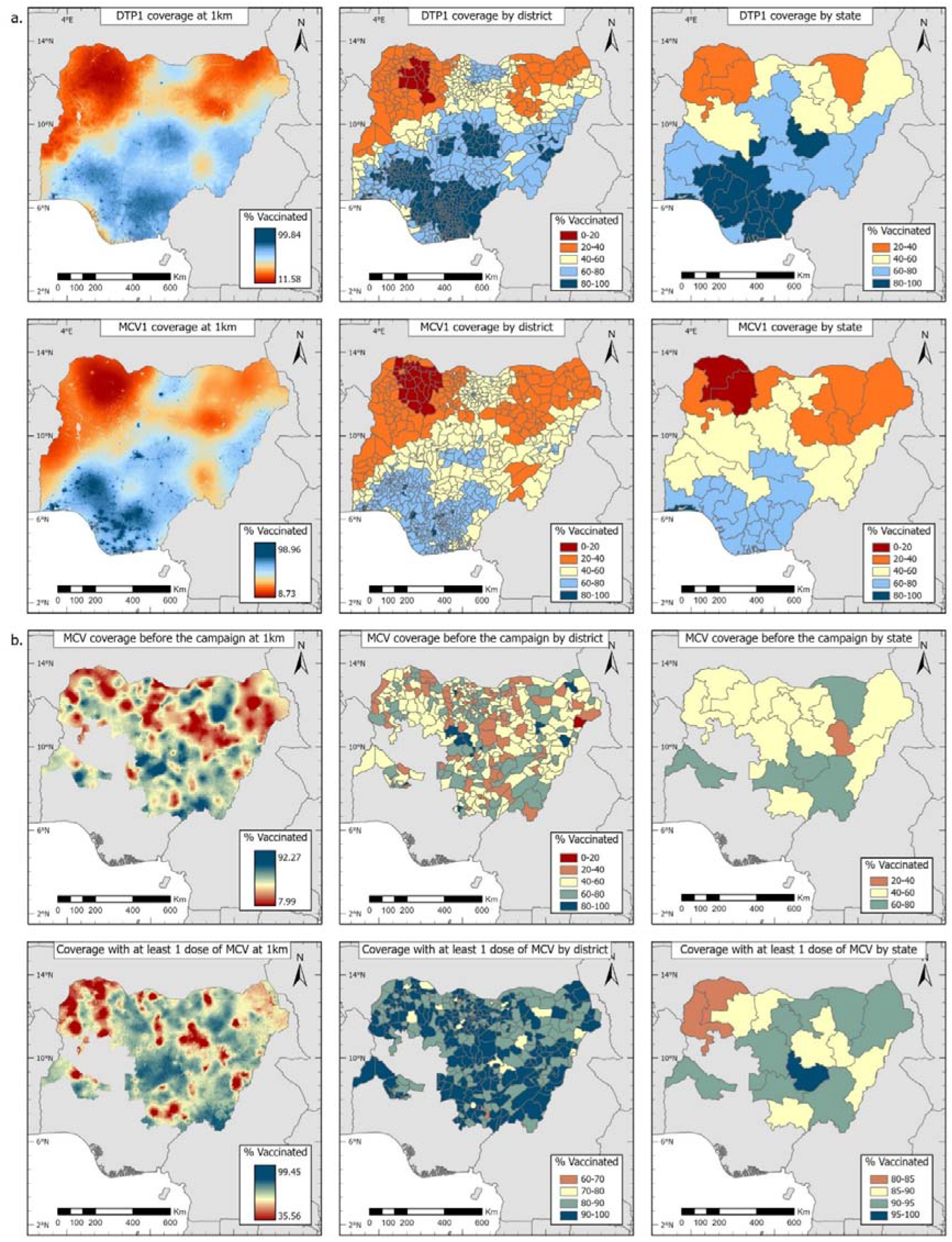
(a) Modelled estimates of crude routine DTP1 and MCV1 coverage for 2018, children aged 12-23 months, 2018 NDHS. (b) MCV coverage before the 2019 campaign and coverage with at least one dose of MCV by the end of the 2019 campaign, children aged 9-35 months, produced using the 2019 PCCS for northern states. Corresponding uncertainty estimates are shown in supplementary Figure 1.

Patterns of MCV coverage before the 2019 measles campaign (Figure 2(b)) were similar to those for the 2018 DHS although in some states, e. g., Sokoto, Taraba, Yobe and Zamfara states, coverage was higher in the former perhaps reflecting recall bias or inclusion of doses received during outbreak response activities. The spatial distribution of MCV1 coverage by the end of the 2019 campaign (coverage with at least one dose of MCV) shows marked improvements in these northern states. However, some heterogeneities still exist as evidenced by the occurrence of pockets of low coverage areas. Notably, some states with lower RI coverage levels in 2018 (e.g., Sokoto and Zamfara) also had among the lowest coverage after the campaign. The patterns in the uncertainties associated with these coverage estimates (supplementary Figure 1) mostly reveal lower precision in areas where data are sparse. These also show that the precision of the estimates increases substantially with decreasing spatial detail.

To enable a fuller assessment of disease risk, we present the estimated distributions of numbers of zero-dose children at the district level in Figure 3 corresponding to the coverage estimates presented earlier (state level zero-dose estimates are shown in supplementary Figure 3). These figures show strong similarities between MCV and DTP zero-dose estimates in 2018, with high-prevalence districts concentrated mostly in the northeastern and northwestern regions, generally mimicking the patterns observed in the coverage maps (Figure 2). However, there are also high-prevalence districts in other areas such as the FCT and the southwestern states of Lagos and Ogun. Districts with the highest DTP zero-dose prevalence (≥ 50,000 unvaccinated children aged under 5 years or under 5s) were in Zamfara, Kebbi and Gombe states; and for MCV, Zamfara, Kebbi, Gombe, FCT, Bauchi, Kaduna and Yobe states. Importantly, for both vaccines, these districts with the highest zero-dose prevalence do not include some of the lowest coverage districts identified earlier (e.g., some districts in Sokoto state), which had low population density, and vice versa.

**Figure 3:**
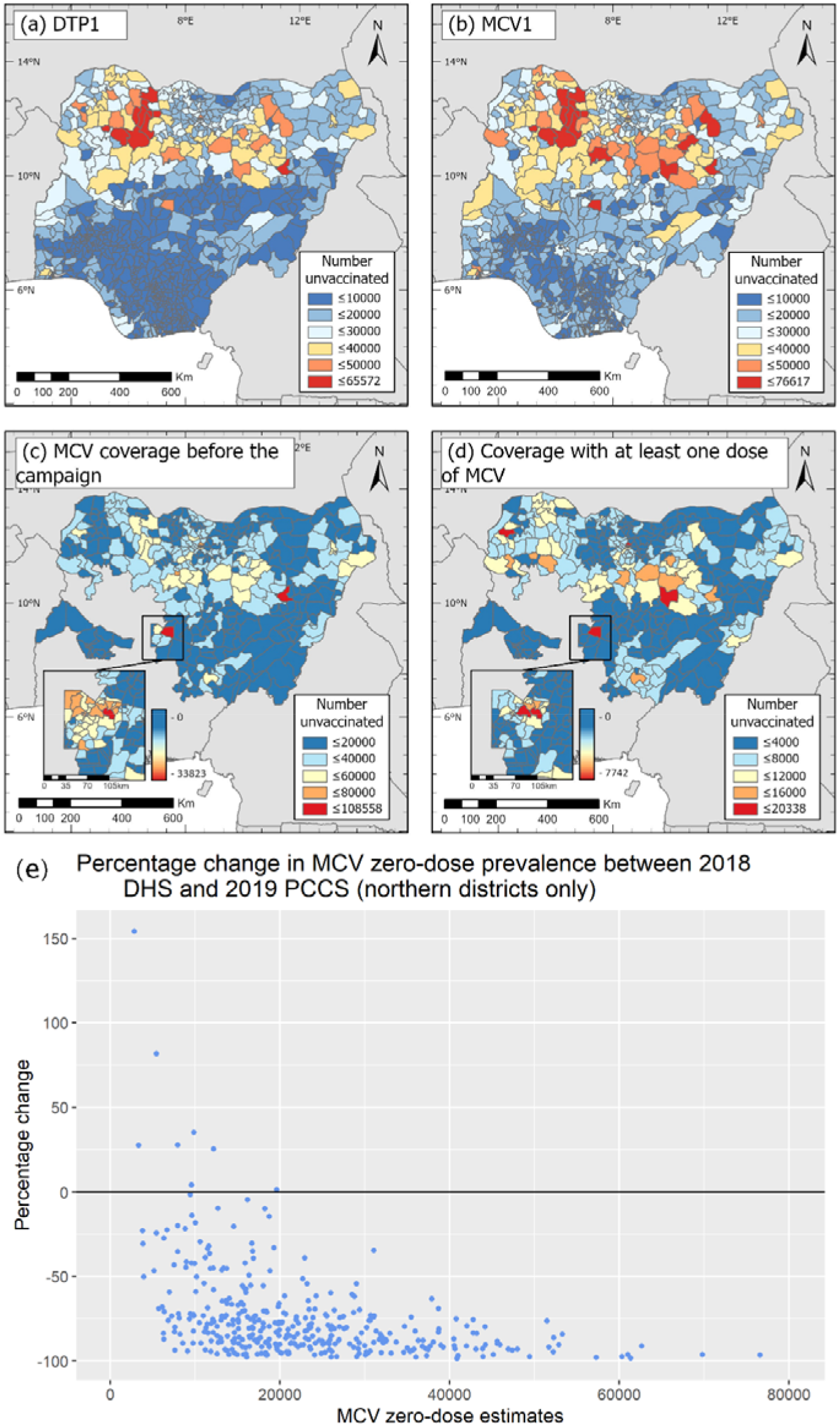
District-level estimates of numbers of zero-dose children among under-5s for (a) DTP and (b) MCV in 2018 (produced using the 2018 NDHS), and MCV in 2019 (c) before and (d) at the end of the 2019 measles campaign (produced using the 2019 PCCS). The insets in panels (c) and (d) show the distributions of zero-dose prevalence at the ward level. Panel (e) shows percentage change in MCV zero-dose prevalence between 2018 and after the 2019 measles campaign in the districts in the northern region.

Figures 3(d) and (e) reveal the extent to which the 2019 measles vaccination campaign had been successful in reducing MCV zero-dose prevalence that had accumulated across the northern states. District-level MCV zero-dose estimates post-campaign are below 20, 338 children as against 76, 617 children prior to the campaign in 2018, revealing large reductions particularly in high zero-dose prevalence districts. However, some areas with relatively high zero-dose prevalence (*>* 16, 000 under 5s) remained after the campaign and these are in Kano (Ugongo), Bauchi (Bauchi), Kebbi (Birnin Kebbi) and FCT (Municipal Area Council). Some districts in Bauchi (e.g., Bauchi, Darazo and Ningi) were consistently high-prevalence districts in 2018 and before and after the campaign. There were also a few districts where it appeared that the campaign was not effective in reaching zero-dose children (Figure 3(e)), although these had lower zero-dose prevalence in 2018 (< 10, 000 zero-dose children).

### Trends in measles incidence and relationships with MCV zero-dose estimates

In Figure 4, we show the spatial distributions of confirmed measles case counts for all age groups at the district and state levels from 2018 to 2020. Our analysis revealed a total of 7,603, 28,440 and 9,394 confirmed cases of measles in the respective years, most of which occurred in children aged under 5 years (see supplementary Figures 4 and 5). The larger number of cases reported in 2019 was due to large measles outbreaks in some districts in Borno state (Maiduguri, Jere and Bama) [30], much of which were either clinically diagnosed or epidemiologically-linked (supplementary Figure 4). There were also spikes in case numbers in some districts in Katsina (Katsina district) in 2018 and 2019.

**Figure 4:**
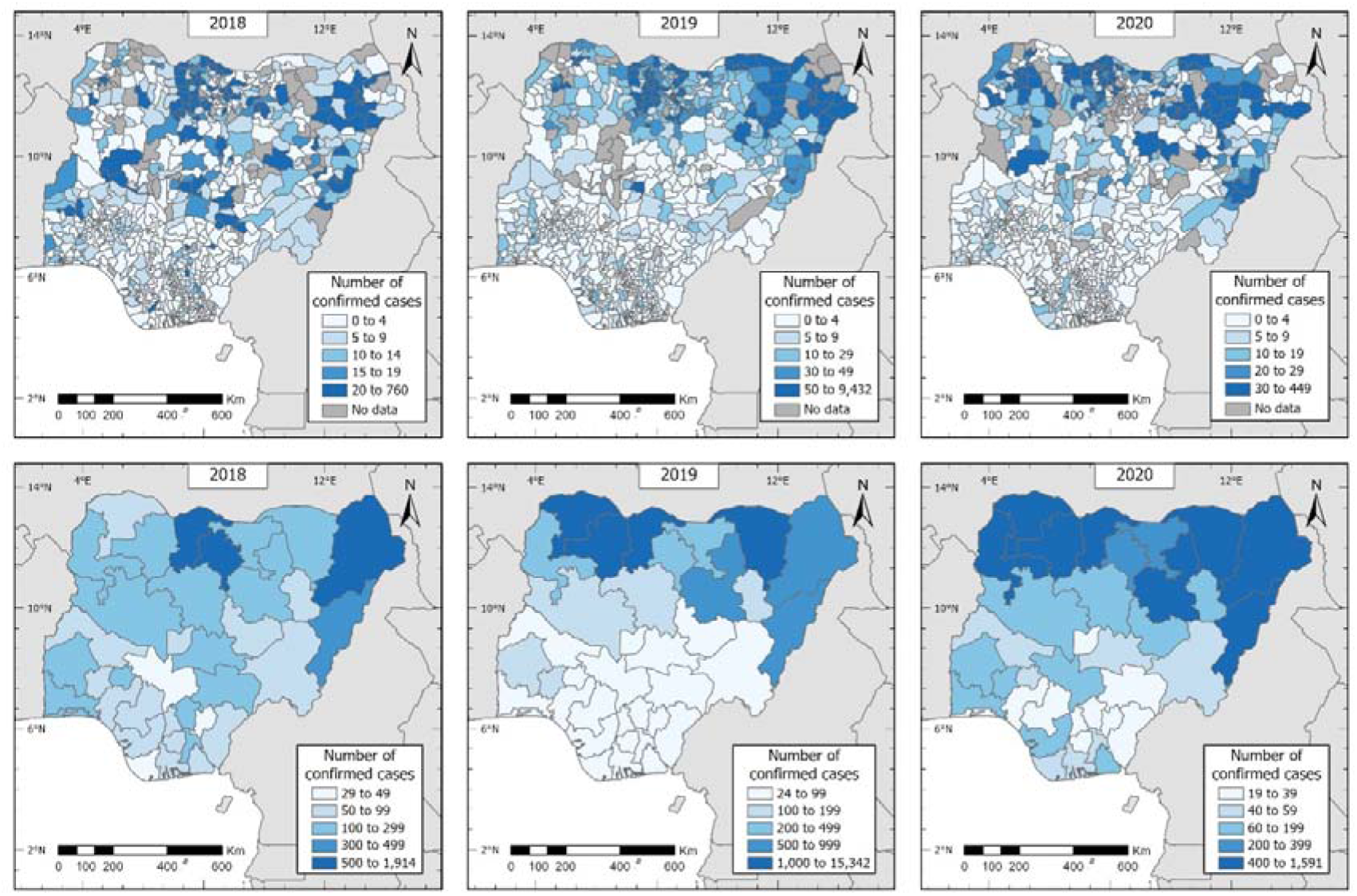
Spatial distribution of confirmed cases of measles in Nigeria reported between 2018 and 2020 at the district (top row) and state (bottom row) levels (all ages).

The distribution of cases according to the number of vaccine doses received (supplementary Figure 6) shows that irrespective of method of diagnosis, most of the confirmed cases occurred in MCV zero-dose individuals, suggesting that poor vaccination coverage rather than decreased vaccine efficacy would have been responsible for these cases. For all three years, the patterns in these maps closely resemble the spatial distribution of RI coverage in 2018 and the corresponding zero-dose estimates (Figures 2 and 3). Notably, there is a marked, persistent north-south divide in case distribution over the years, with a concentration of higher case numbers in districts in the north. The same pattern can also be seen in the measles incidence rates shown in supplementary Figure 7, although there are some minor differences.

At the state level (Figure 5), we estimated correlations of 0.55, 0.60 and 0.57 (Spearman’s correlation coefficient, excluding the outlying observations) between the case counts and MCV zero-dose estimates in 2018 (nationwide) and 2019 (northern states only) – before and after the campaign, respectively. These show strong relationships between measles incidence and the zero-dose estimates obtained through using MCV1 RI coverage and a combination of RI and campaign coverage. In 2018, areas with a combination of higher case counts (between 154 and 1,914 individuals) and higher zero-dose estimates (between 461,091 and 954,963 under 5s) were concentrated in the northern states of Borno, Bauchi, Jigawa, Kano, Katsina, Kaduna, Niger and Kebbi, and the southwestern state of Oyo. In 2019 before the campaign, these areas were in Kano, Kaduna, Katsina, Borno and Sokoto states (zero dose estimate range: 461,557 - 1,146,982 under 5s; case count range: 356 - 15,432 individuals); while after the campaign, these were in Kano, Katsina, Sokoto and Kebbi states (zero-dose estimate range: 116,633 - 230,449 under 5s). Thus, while the campaign was successful in reducing the numbers of zero-dose children in the northern states as demonstrated earlier, considerable numbers of under 5s remained unvaccinated in some areas. We note that the reported case count for Sokoto state was lower in 2018 even though it had a higher zero-dose estimate for the same year.

**Figure 5:**
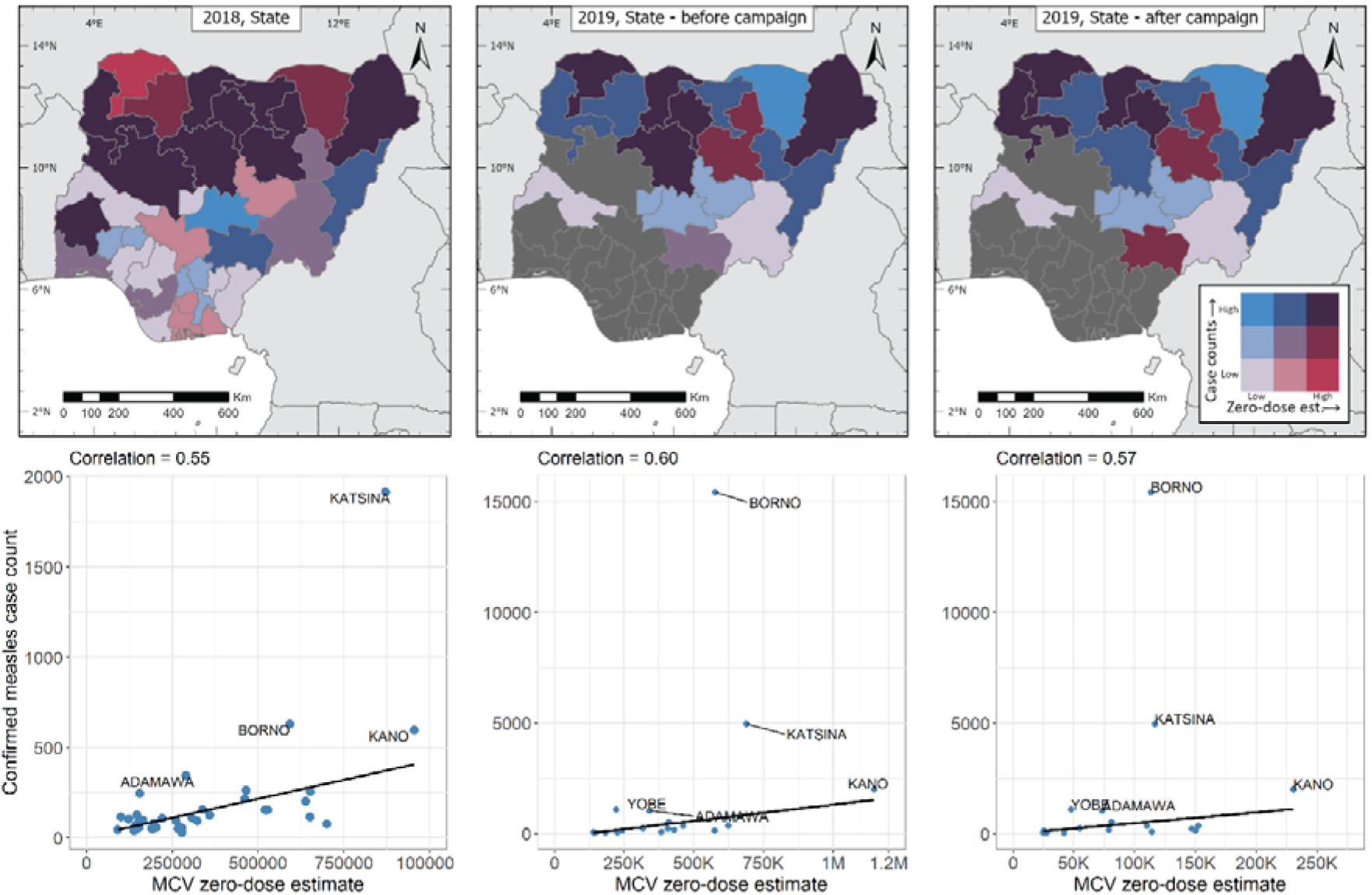
Top row: Joint spatial distribution of confirmed measles case counts (all ages, in blue) and MCV zero-dose estimates (in red) at the state level in 2018 and 2019. Bottom row: Plots showing the relationships between confirmed measles case counts (all ages) and MCV zero-dose estimates in 2018 and 2019 at the state level. The black lines are simple least square fits to the data.

At the district level (supplementary Figure 8), the estimated correlations were poorer for both years (0.36 in 2018, and 0.29 (before campaign) and 0.20 (after campaign) in 2019), with most of the cases occurring in districts where the zero-dose estimates fall between 10,000 and 50,000 under 5s for RI coverage and ≤ 10,000 under 5s for campaign coverage.

In Figure 6, we compare the spatial distribution of measles case counts in 2020 with MCV zero-dose estimates in 2018 due to lack of survey coverage estimates in 2020 at the time of analysis. First, we observe that despite the measles vaccination campaign in the northern states in 2019, the spatial distribution of the case counts in 2020 reveals higher incidence in the north (Figure 4) and this correlates strongly with the distribution of cases in 2018 (correlation = 0.61 at the state level and 0.39 at the district level), suggesting a replay of the 2018 scenario. In particular, all the states designated as high risk by virtue of higher zero-dose estimates with or without higher confirmed case counts, by the end of the campaign in 2019 (except Nasarawa state), had higher numbers (≥ 200) of confirmed cases in 2020. Further, we estimated correlations of 0.82 and 0.42 between the case counts in 2020 and zero-dose estimates in 2018 at the state and district levels respectively, which points to the failure of the routine immunization system in these areas to maintain the gains achieved via campaigns. We also observe that almost all the northeastern and northwestern states (except Gombe, Adamawa and Taraba) and Niger state had a combination of higher case counts (between 196 and 1,591 confirmed cases) and higher zero dose estimates. At the district level, there are visible clusters of high-risk areas (i.e., areas with a combination of higher case counts and higher zero-dose estimates) within the high-risk states.

**Figure 6:**
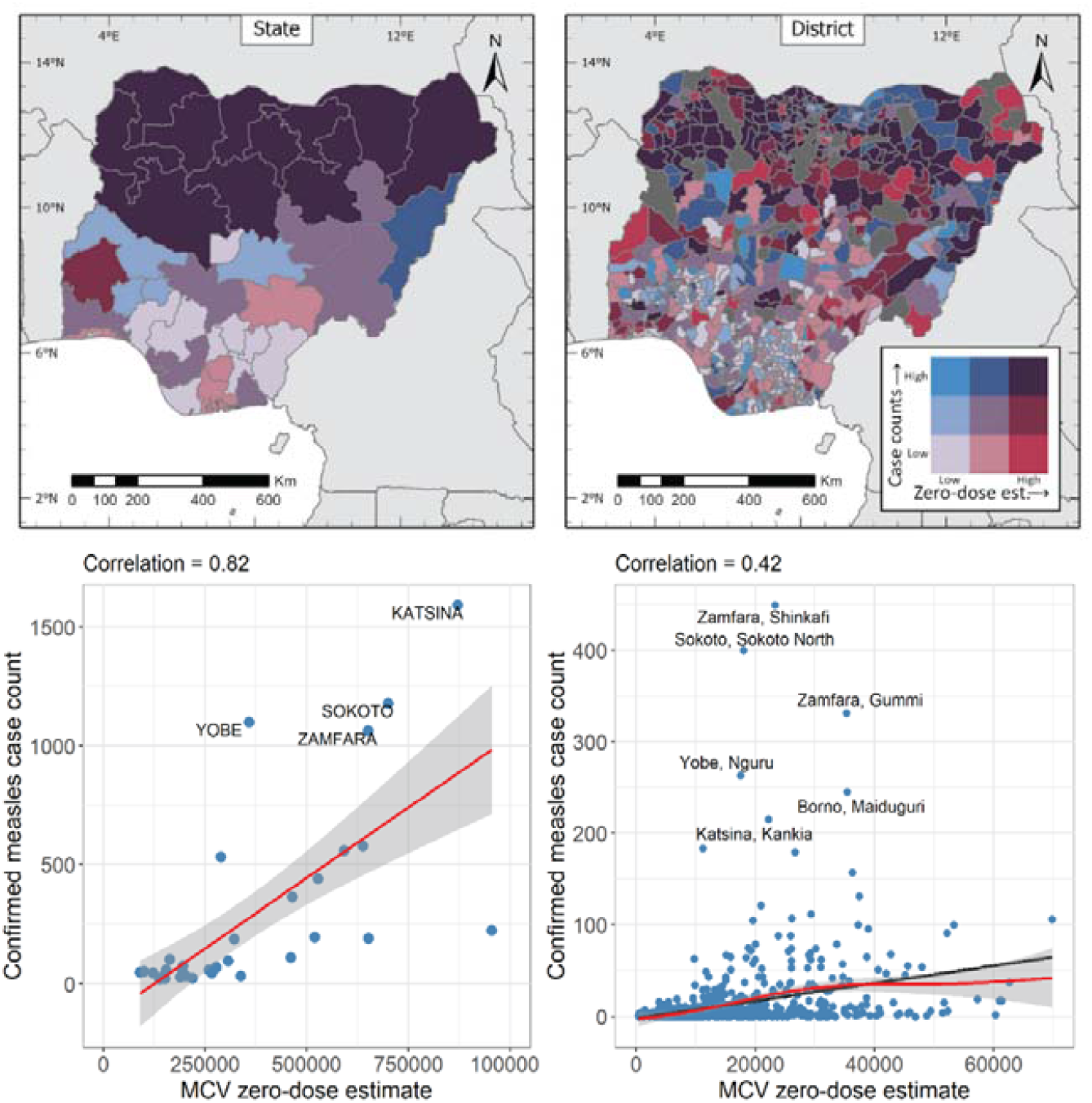
Relationships between confirmed measles cases in 2020 and MCV zero-dose estimates in 2018 at the state (left panels) and district (right panels) levels. In the bottom panels, the red lines and grey coloured bands are natural splines fits and corresponding uncertainty intervals while the black lines are simple least square fits to the data.

## Discussion

Immunization Agenda 2030 [12] has a target of reducing the number of zero-dose children substantially by reaching high and equitable coverage levels by 2030. To interrupt measles transmission and prevent outbreaks, this means reaching at least 95% coverage with both recommended doses of MCV. Our analyses revealed that Nigeria is far from reaching these targets due to suboptimal RI performance and the failure of frequent campaigns to achieve or maintain such high coverage levels, resulting in the persistence of highly heterogeneous coverage levels across the country. The lower coverage in northern states is especially concerning for measles transmission [31] because these states also have higher birth rates [20, 32], which facilitates transmission as well as increasing the challenge of sustaining RI coverage for a generally weaker health system and infrastructure [33].

Our work demonstrates the utility of geospatial analysis for uncovering areas with suboptimal routine immunization systems and where campaigns have been less effective in terms of reaching previously unvaccinated children, both of which are crucial for designing effective strategies to reach missed communities and high-risk areas. We found that districts where RI performance had been significantly lower and which had among the highest zero-dose prevalence were mostly located in the northeastern and northwestern parts of the country, although some high RI zero-dose prevalence districts were found in other parts of the country which had high population densities, e.g., Lagos and Ogun states. We also estimated that some of these districts (e.g., Municipal Area Council in the FCT and Bauchi in Bauchi state) had some of the highest residual MCV zero-dose prevalence after the 2019 measles vaccination campaign, although the campaign was shown to reduce pre-campaign MCV zero-dose prevalence substantially. The persistence of these clusters of vulnerabilities pose significant risk to disease control [8]. Hence, the estimation of residual zero-dose prevalence following vaccination campaigns should be analysed and reported by PCCS and can contribute to the design of follow-up activities. Unfortunately, this has been done infrequently to date and in countries with higher RI coverage than Nigeria, it can be difficult and costly to undertake PCCS with the goal of estimating coverage among previously unreached children [34].

Our analyses further revealed that despite SIAs reducing predicted measles incidence in Nigeria [35], they have not interrupted transmission and incidence continues to be related to routine immunization coverage. For both non-campaign years that we analysed – 2018 and 2020, we found strong correlations between confirmed measles case counts and MCV zero-dose estimates. In 2019, we also observed strong correspondence between the case counts and the zero-dose estimates, both before and after the 2019 measles campaign in the northern states. Although the SIA substantially improved coverage levels relative to RI, only Plateau state and very few districts achieved ≥ 95% coverage, and 147 districts (out of 373) had below 90% coverage. Furthermore, the PCCS (like DHS) omitted the most insecure areas from the sampling frame and a small number of selected clusters in the survey could not be visited due to conflict. Areas omitted are likely to have lower RI and SIA coverage [36, 37]. Thus, it is not surprising that the campaign had not been fully effective in controlling the spread of measles, especially in the north. Gains from SIAs are shorter-lived in northern than southern states, due to the higher birth rate and lower RI coverage in the north. Recognising this, Nigeria scheduled SIAs in the north 2 years after the previous SIA while an SIA was planned in 2020 in the south but postponed due to the COVID-19 pandemic. While RI coverage is so low in the north, however, an annual SIA in northeast and northwestern states could be a more effective strategy, though more costly [35]. Alternatively, where there is evidence that frequent SIAs reach more previously vaccinated children, then the implementation of targeted SIAs or RI interventions such as multi-antigen periodic intensification of routine immunization in districts with higher zero-dose prevalence could be an even more effective option for disease control. Our analyses also revealed strong similarities between the spatial distributions of MCV and DTP zero-dose estimates in 2018, further pointing to weaker RI systems in the northern parts of the country.

We found that the dropout rates between the routine coverage of DTP1 and MCV1 (relative to DTP1 coverage) in the 2018 NDHS varied substantially across the country. Some areas with the highest (positive) dropout rates such as Akwa Ibom, Cross-River and Plateau states also had higher DTP1 coverage, suggesting that factors other than access to vaccination services may have been responsible for the low uptake of MCV1 in these areas – see Aheto et al [18]. In the northeast and northwest, where DTP1 coverage was lowest, some areas had higher MCV1 coverage which may reflect inadvertent classification of SIA or outbreak response campaign doses as RI doses for MCV. Vaccination records were only seen for 40% of children in the 2018 NDHS (and only 33.9% and 28.5% in northeast and northwest regions, respectively), hence reliance was placed on the mother’s ability to correctly state whether the child had received MCV and to distinguish between routine and campaign doses. Importantly, others have found that opportunities to administer MCV1 when eligible children attend health facilities for other vaccines are more frequently missed in northern states than southern, although missed opportunities for simultaneous vaccination have decreased over time [38]. These findings point to the need for regional or subnational approaches when investigating/analysing the drivers of vaccination coverage in the country. Such analyses are also likely to be beneficial for designing effective strategies to reach zero-dose children.

Our work revealed stronger correlations between MCV zero-dose estimates and measles case counts at the state level compared to the district level. This may be due to data quality issues such as diagnosis occurring outside a patient’s district of residence, non-reporting of cases, likely variation in the accuracy of clinical diagnosis of measles by time and place which may have affected the distribution of the reported case counts at the district level. Also, the uncertainties associated with coverage estimates underlying the zero-dose estimates are generally higher at the district level than the state level. Hence, although the importance of spatially detailed estimates of health and demographic indicators for health policy and decision-making and resource allocation is well recognized, there is a need for greater investments to boost the quality of data available at smaller geographical units.

Our analyses are subject to some limitations. Our vaccination coverage estimates are based on information obtained from vaccination cards as well as via caregiver recall and are hence subject to information/recall bias [39]. The sampling frames used for the surveys analysed may have missed important hard-to-reach/disadvantaged populations such as those living in conflict areas or urban slums. This may have led to an underestimation of the zero-dose prevalence in some areas. The estimation of vaccination coverage and associated zero-dose prevalence for these at-risk populations can be much improved in future analysis through using more accurate data from targeted surveys. Geospatial data from the 2018 NDHS and 2019 PCCS analyzed in our work were based on displaced geographical coordinates at the cluster level. Although these displacements do not generally result in the coordinates being positioned outside their districts of origin, it is likely that these may have had some effect on the coverage estimates and resulting zero-dose estimates particularly at more granular levels where clear distinctions between types of residence (formal urban, urban slums and rural settlements) may be required [40]. The accuracy of the zero-dose estimates presented in our work depends largely on the accuracies of the underlying population and coverage estimates. We did not account for the uncertainties in the population (and coverage) estimates [41, 42] when producing the zero-dose estimates. In practice, this could be best done using a joint modelling approach which is beyond the scope of our work. We did not obtain an official endorsement from the Nigerian government to use WorldPop data for producing the zero-dose estimates, although these data have been used widely in similar contexts [22, 43]. Also, by using the distribution of zero-dose children to assess the performance of vaccine delivery strategies, our study precludes scenarios where certain demand-side barriers to immunization cannot be overcome through effective and efficient immunization service delivery. Our study did not assess the timeliness of vaccination vis-à-vis the distribution of zero-dose children, considering that these children are right-censored in our analysis and may be vaccinated at a later time. Nevertheless, the best framework to evaluate the effect of timeliness of vaccination on zero-dose prevalence is a longitudinal study.

Furthermore, the measles case-based surveillance data analysed here are an underestimate of measles incidence, since most cases may not have been reported [16]. Only a small proportion (16.4% overall; and 22.8%, 11.3% and 27% for 2018, 2019 and 2020, respectively) of the confirmed cases included in our study were confirmed by a laboratory, signalling additional data quality issues. Stockouts of measles laboratory test kits, loss of accreditation in 2018 of the measles regional reference laboratory in Gombe, staffing shortages and sample transportation challenges have contributed to poor measles serum sample testing rates. Overall, VPD surveillance was additionally impacted during 2020 by the COVID-19 pandemic, particularly due to significant shifts of resources and priorities from VPD surveillance to COVID-19 response, and lockdowns and bans on interstate movement reducing access to health facilities and uptake of health services, and hindering sample transport. Also, the preponderance of missing age information in the data hindered our ability to undertake any meaningful age-dependent analysis of the confirmed cases. Hence, we undertook comparisons of all-age case counts with the zero-dose estimates. We did not investigate the role of migration (e.g., rural-urban migration) and conflict on measles incidence. However, we note that the spike in case counts observed in Borno state in 2019 (Figure 5) is likely due to the disruption to vaccination services caused by insurgency. Also, higher confirmed case counts recorded in urban districts (e.g., Maiduguri and Katsina districts – see supplementary Figure 8) may have been due to rural-urban migration, including those fleeing to the urban areas due to high spate of insurgency within the mostly remote rural areas, as well as other contributory factors such as better surveillance and slum areas [44]. Lastly, the stark differences in estimated MCV RI coverage from the 2018 DHS and the pre-campaign data recorded in the 2019 PCCS calls for improvements in data quality, particularly improved retention of up-to-date HBRs from which more accurate data can be obtained.

Nigeria has committed to a target of 30% reduction in the number of zero-dose children by 2025. The estimated number of routine zero-dose children could be an indication of health system performance and a proxy measure for broader health outcomes of communities that have clusters of zero-dose children. Further analyses categorizing the estimated distribution of zero-dose children as unreached (programme delivery failure), far-to-reach (distance, easily solvable through additional mobility support), hard-to-reach (insecurity, difficult terrain), and never reached (unmapped, unknown), could help with the selection and resourcing of appropriate programmatic interventions. Nigeria is currently developing a zero-dose and unreached strategy for routine immunization which is building on the above categorization and could become a part of its broader health and immunization strategy. During the recent SIAs, a zero-dose reduction operation plan (zero-drop initiative) was launched by the involvement of supplementary immunization officers and supporting them to reach more unreached settlements and zero-dose children. The challenges to VPD surveillance due to COVID-19 pandemic described previously also impacted RI coverage in Nigeria during 2020-22. COVID-19 vaccine hesitancy affected the uptake of routine vaccines, due to fear that a COVID-19 vaccine would be administered instead of or in addition to other antigens being offered during RI or SIAs. The nationwide measles vaccination campaign that had been planned for early 2021 was postponed due to the pandemic and global supply shortage. Only 13 northern states implemented the campaign during the fourth quarter of 2021, and implementation in the remaining states was postponed to June 2022 in three states and September-October 2022 in 21 states. Combined with the reduction in RI coverage, the delayed campaigns resulted in a build-up of a large cohort of susceptible children and increased measles outbreaks and related deaths [45]. Also, economic impacts arising from the pandemic and other factors have led to an increase in insecurity and insurgency in Nigeria, including areas not previously affected, further impacting vaccination coverage.

In future work, we will explore the utility of the zero-dose estimates for optimizing the placement and improvement of vaccination posts both for outreach RI activities and SIAs, as well as integration with health facility catchments to facilitate the design and implementation of localized interventions to improve vaccination services. We will also explore alternative definitions of “zero dose” such as non-receipt of any of the four basic vaccines (bacille Calmette-Guérin vaccine (BCG), DTP, oral polio vaccine (OPV) and MCV). Although our work has revealed interesting similarities between MCV and DTP zero-dose, it will be informative to understand how these compare with the spatial distribution of children who had not received any of the basic vaccines. We will conduct multi-level analyses similar to Aheto et al [18] and Utazi et al [37], but at the regional level, to better understand regional differences in the major drivers of poor vaccine uptake. Beyond the descriptive analysis using measles case-based surveillance data presented here, we will explore different options to model and refine the data using geospatial approaches. These data can also be incorporated into geostatistical models of vaccination coverage in a fusion modelling framework to improve coverage estimation. We will also examine how the spatial distribution and frequency of measles outbreaks (and those of other diseases such as yellow fever, cholera and circulating vaccine-derived polio virus), as against the case counts used in this work, relate to the distribution of zero-dose children, and also possibly develop a framework for predicting outbreaks. This will be useful for understanding where health systems require strengthening. We will consider developing an online data visualization tool to facilitate access to and utility of the outputs of the current work and other future work by policy makers and other researchers. Finally, we will seek an expansion of the work presented here and other follow-on analyses to other countries.

## Data Availability

All data used in the work are publicly available via the sources referenced in the manuscript.

## Acknowledgements

This work was supported by funding from the Bill & Melinda Gates Foundation (Investment ID INV-003287).

## Competing interests

The authors declare no competing interests.

## Notes

### Competing Interest Statement

The authors have declared no competing interest.

