## supplementary for "Mapping the distribution of zero-dose children to assess the performance of vaccine delivery strategies and their relationships with measles incidence in Nigeria"

**Supplementary file**

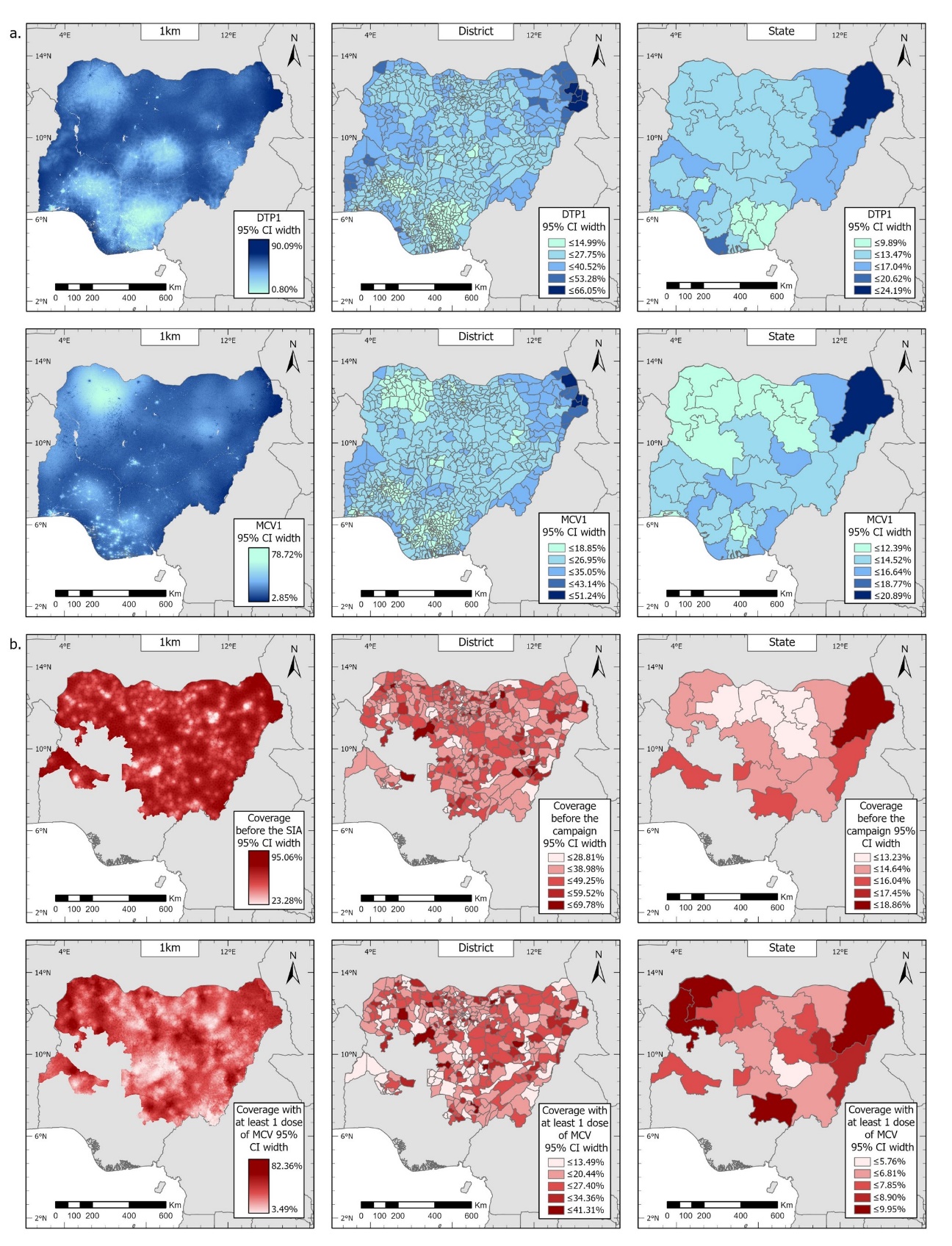

Figure 1: (a) Uncertainties associated with modelled estimates of routine DTP1 and MCV1 coverage for 2018, shown as the widths of the corresponding 95% credible intervals. (b) Uncertainties associated with modelled estimates of MCV coverage before the 2019 campaign and coverage with at least one dose of MCV by the end of the 2019 campaign, shown as the widths of the corresponding 95% credible intervals.

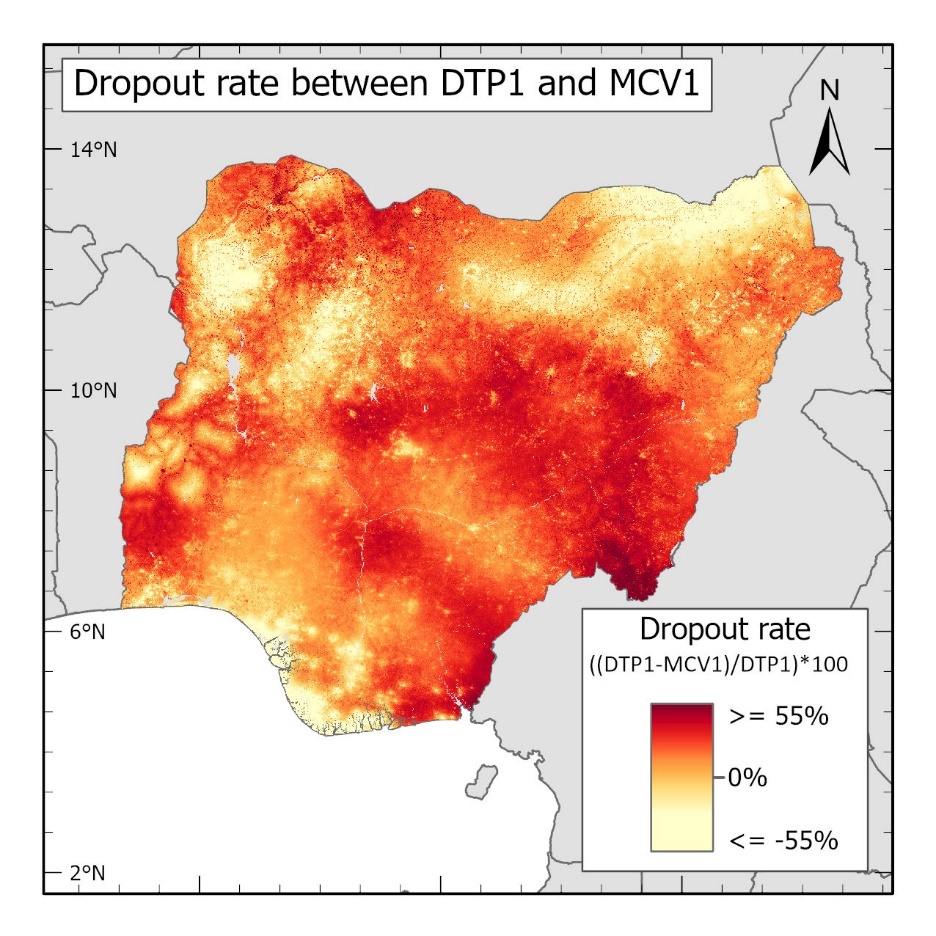

Figure 2: Dropout rate between modelled estimates of routine DTP1 and MCV1 coverage in 2018.

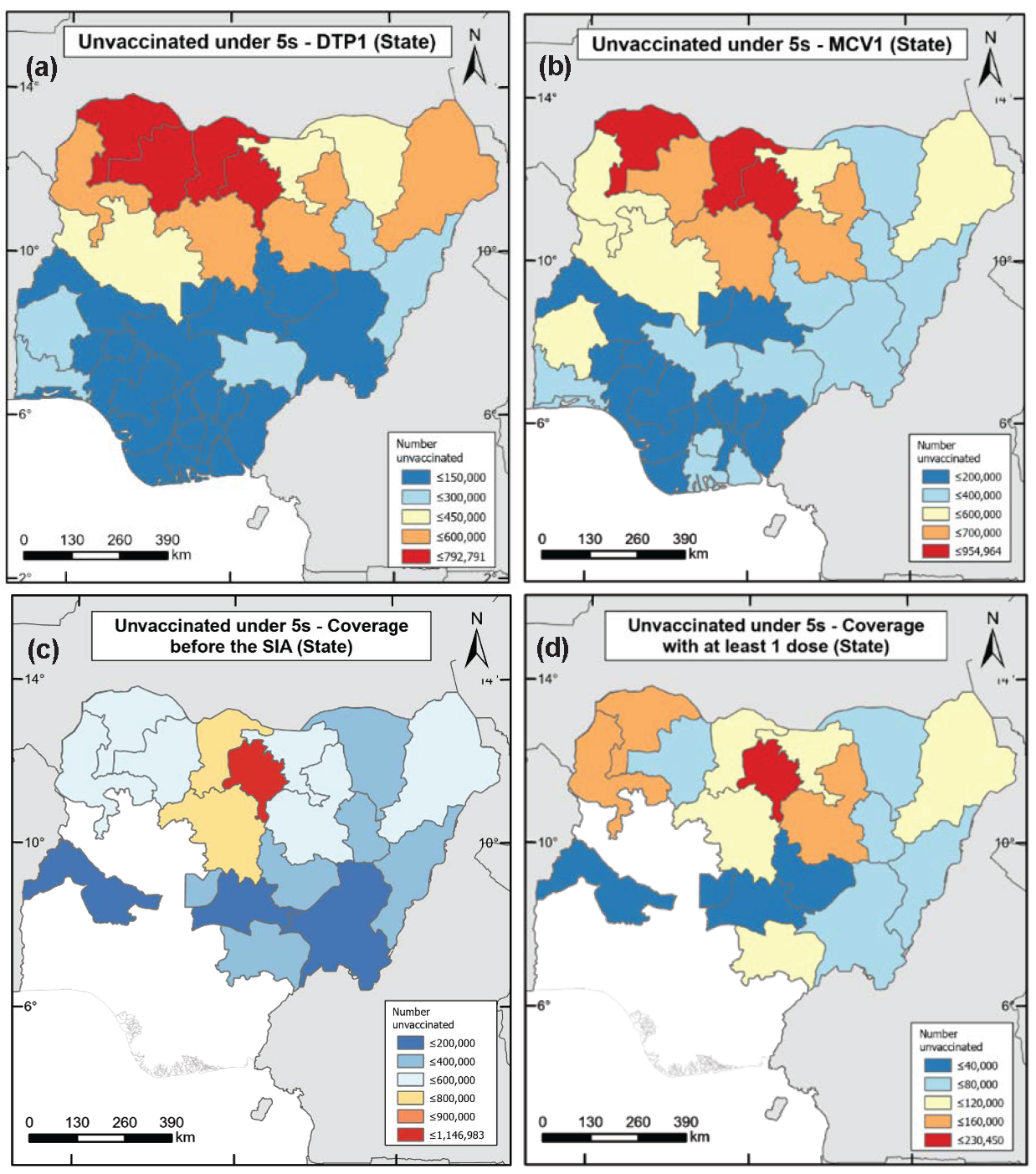

Figure 3: Estimates of numbers of zero-dose children among under-5s for (a) DTP1 and (b) MCV1 in 2018 (produced using the 2018 DHS), and MCV1 in 2019 (c) before and (d) at the end of the 2019 measles campaign (produced using the 2019 PCCS).

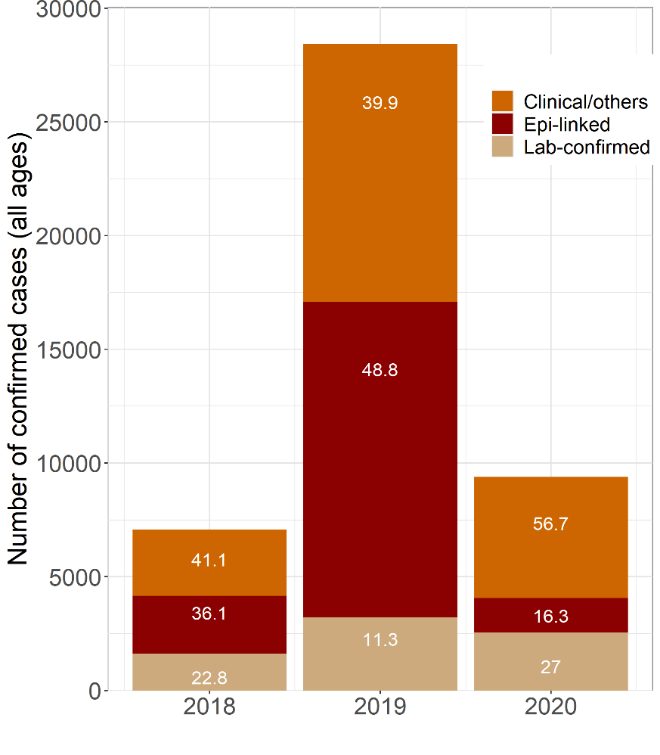

Figure 4: Distribution of confirmed measles case counts in Nigeria between 2018 and 2020 according to method of diagnosis.

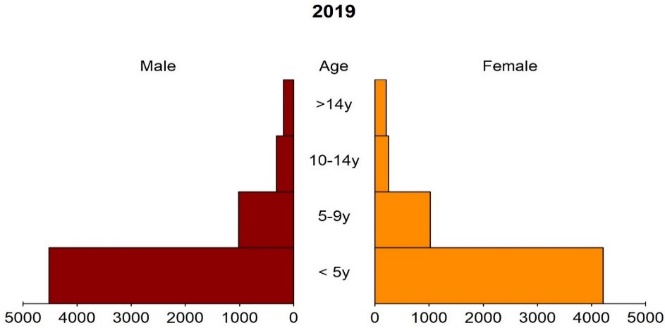

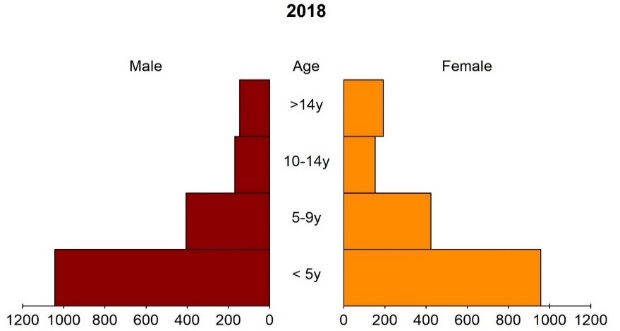

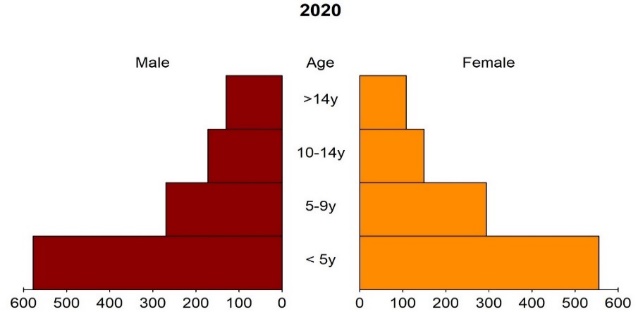

Figure 5: Age-sex distribution of confirmed measles case counts in Nigeria between 2018 and 2020.

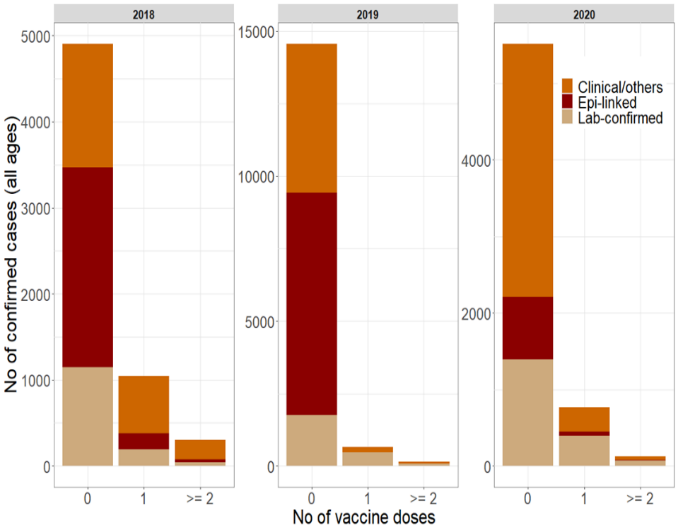

Figure 6: Distribution of confirmed measles case counts by method of diagnosis and number of vaccine doses received.

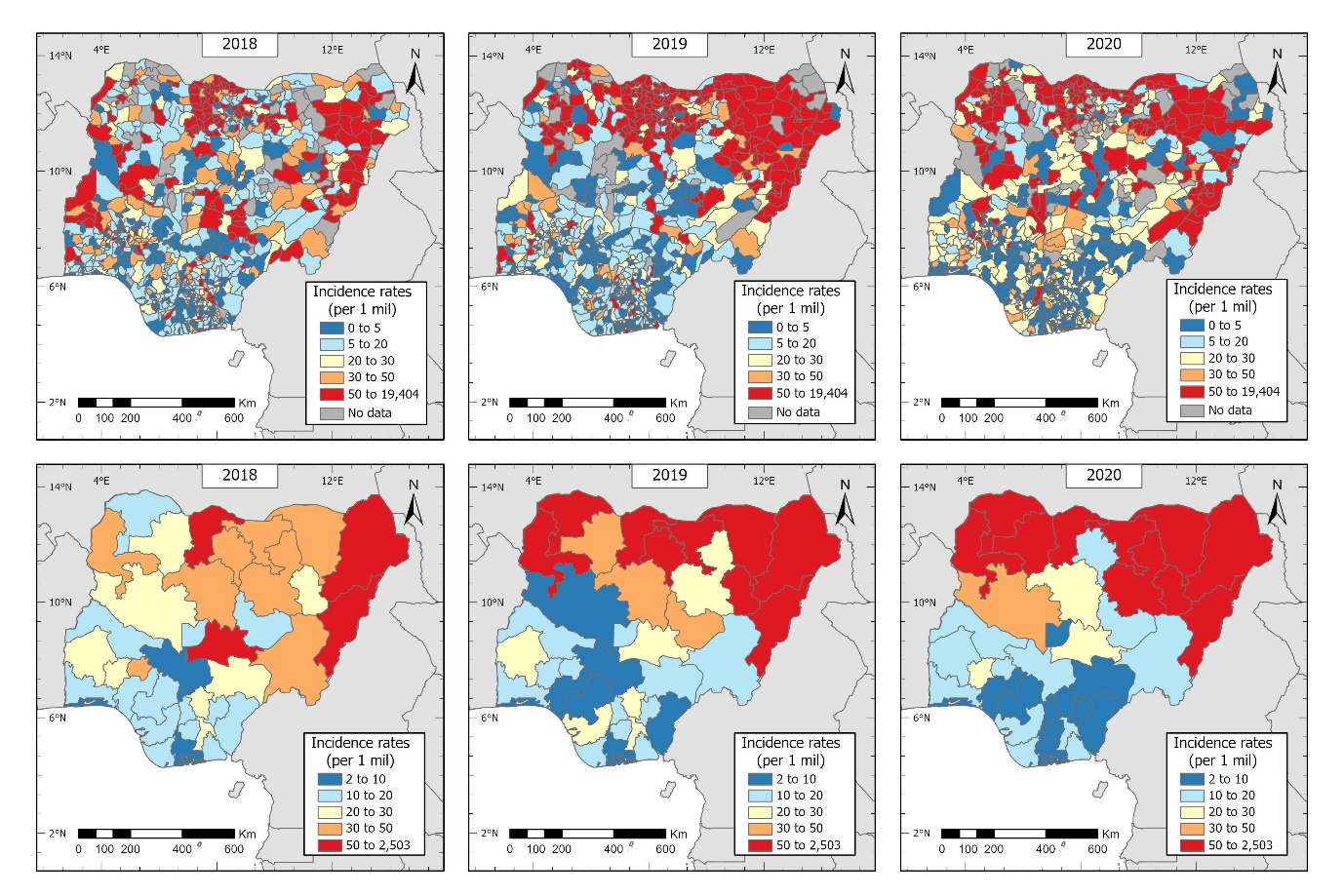

Figure 7: Measles incidence rates in Nigeria between 2018 and 2020 at the district (top) and state (bottom) levels.

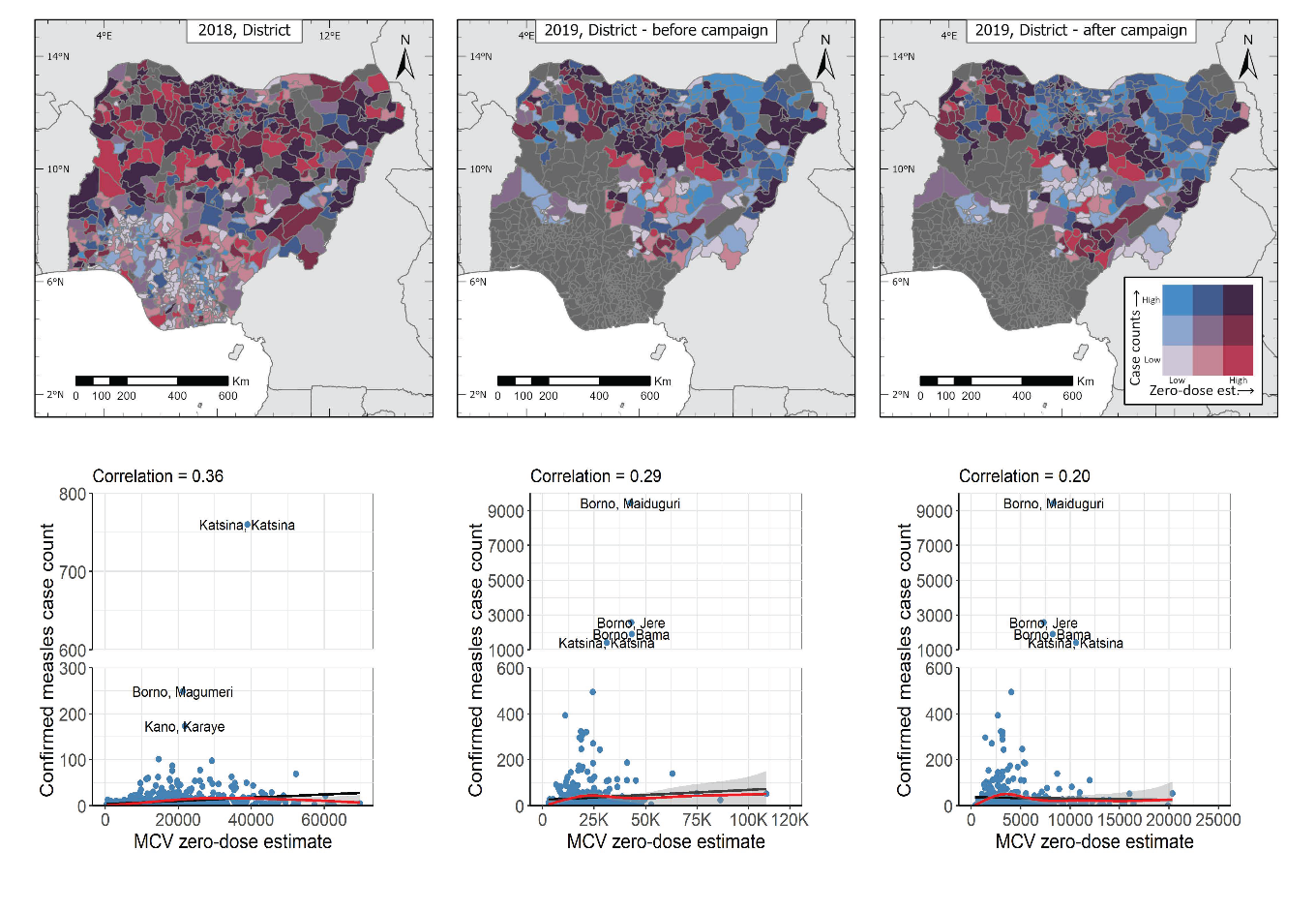

Figure 8: Top row: Joint spatial distribution of confirmed measles case counts (all ages, in blue) and MCV zero-dose estimates (in red) at the district level in 2018 and 2019. Bottom row: Plots showing the relationships between confirmed measles case counts (all ages) and MCV zero-dose estimates in 2018 and 2019 at the district level. The black lines are simple least square fits to the data while the red lines and grey coloured bands are natural splines fits and corresponding uncertainty intervals.
